# EASing Oxytocin in Early Labour - OUTcomes for Mothers and Babies (EASE-OUT): a feasibility randomised controlled trial

**DOI:** 10.64898/2026.07.18.26358404

**Authors:** Alanna Krisanaleela, Belinda R. Bruce, Hala Phipps, Rachel Morton, Jon Hyett, William Tarnow-Mordi, Adrienne Gordon, Shideh Pakzadian, Karen Lawrence, Melissa Wang, Bradley S. de Vries

## Abstract

**Objective:** To assess the feasibility of a definitive randomised controlled trial comparing halved versus routine-concentration oxytocin infusion during active phase of the first stage of labour in women undergoing induction of labour.

**Design:** Multicentre, double-blind, randomised feasibility trial with 1:1 allocation.

**Participants:** Women & birthing people aged 18 years or older undergoing planned induction of labour in two public maternity hospitals in Sydney, Australia, between September 2023 and September 2025 were eligible. Key exclusions included previous caesarean delivery, pre-existing diabetes, major fetal anomaly, abnormal fetal cardiotocography, malpresentation, suspected cephalopelvic disproportion, suspected chorioamnionitis, intrapartum pyrexia, and other predefined maternal or fetal safety concerns.

**Interventions:** All participants commenced induction with standard oxytocin during the latent phase (10 IU in 1 L crystalloid). At established active labour, participants were randomised to receive either halved-concentration oxytocin (5 IU in 1 L crystalloid) or routine-concentration oxytocin (10 IU in 1 L normal saline), titrated according to the New South Wales (NSW) Health oxytocin protocol between one and 40 mIUmin^-1^.

**Main outcome measures:** Feasibility outcomes were recruitment, consent, randomisation, retention, protocol adherence, unblinding, and treatment separation, assessed by total oxytocin dose and infusion rates. Secondary outcomes included maternal, neonatal, and participant-reported outcomes.

**Results:** Of 771 women assessed for eligibility in two centres, 572 were approached and 293 (51%) consented to participate, of whom 101 (34%) were randomised. During active recruitment, randomised participants represented 9.2% of all births. Baseline characteristics were similar between groups. No between-group differences were observed in maternal, neonatal, or participant-reported outcomes, although the trial was not powered to assess clinical effectiveness. Satisfaction outcomes were ascertained for 69% (70/101) participants. There was 100% ascertainment for short-term clinical outcomes and for readmissions to the hospital of delivery. Twenty-nine participants were interested in providing feedback and in being involved in developing a larger study across multiple hospitals.

**Conclusions:** A blinded randomised controlled trial of oxytocin dose reduction in active labour is feasible in a real-world labour ward.

**Trial registration:** This trial was registered on the ANZCTR (ACTRN12622001342707)

**Funding:** This trial was supported through seed funding from the Sydney Institute for Women, Children and their Families, Sydney Local Health District and the NHMRC Clinical Trials Centre, University of Sydney.

## Introduction

In Australia, the proportion of women having labour induced has risen over the past two decades. In 2023, one in three women had their labours induced including 40% of all nulliparous women.^1^ Similar international trends have been reported, with indications broadening over time.^2-4^

Most inductions of labour include the administration of oxytocin, a synthetic hormone analogous to the naturally occurring peptide hormone that acts on the uterus to stimulate smooth muscle contraction during labour.^1^ Oxytocin infusions are titrated to create uterine contractions of sufficient strength and frequency to lead to a vaginal birth, but too much oxytocin can lead to uterine hyperstimulation and consequently fetal hypoxia/asphyxia (‘fetal distress’) that can increase the risk of neonatal encephalopathy and serious long-term disability.^5-7^ Despite extensive use worldwide, no consensus exists regarding the optimal administration of oxytocin. Given the association between hyperstimulation and adverse maternal and neonatal outcomes, interventions to reduce hyperstimulation could improve these outcomes.

A systematic review of discontinuing oxytocin in the active phase of labour identified a reduction in fetal heart rate abnormalities, uterine tachysystole and the risk of caesarean delivery, but an increase in the mean length of labour by 30 minutes.^8^ However, there are significant barriers to implementation of this intervention, and the oxytocin needs to be restarted in more than a third of cases.^8,9^ A Cochrane systematic review comparing high and low-dose oxytocin regimes from the onset of induction of labour found increased rates of fetal distress in high dose regimens but overall, no difference in rates of operative delivery.^10^

We performed this feasibility study to determine whether a randomised controlled trial (RCT) evaluating halving the oxytocin dose during the active phase of labour would be an appropriate and feasible trial design, particularly with respect to i) adherence to protocol and ii) maternal recruitment and retention.

## Materials and Methods

### Trial design

The EASE-OUT feasibility study was a multicentre, double-blinded, randomised controlled feasibility trial, with an allocation ratio of 1:1, undertaken across two major hospitals in Sydney, Australia, over a two-year period from September 2023 to September 2025. The study protocol underwent review and received formal approval from the Ethics Review Committee of the Sydney Local Health District prior to commencement of participant recruitment (Approval No. X22-0294).

All individuals enrolled in the study provided written informed consent. Potential participants were approached in person by a research midwife or trial investigator trained in consent procedures and provided with verbal information about the study, including its aims, procedures, potential risks, and anticipated benefits. Following this discussion and an opportunity to ask questions, participants were given a participant information sheet, available electronically via QR code and in hard copy format. Eligible participants who subsequently provided electronic informed consent underwent eligibility screening by a research midwife or trial investigator trained in study procedures to confirm suitability for enrolment. Randomisation occurred only after informed consent has been obtained and ongoing eligibility had been confirmed.

Oversight of participant safety and study integrity was supported by an independent Data Safety Monitoring Board (DSMB), which regularly reviewed accumulating trial data. The DSMB evaluated all reported serious maternal and neonatal complications and ensured that any emerging safety concerns were promptly identified and appropriately managed.

There were no changes to the methods or the protocol after randomisation commenced.

### Participants

Eligible participants were nulliparous or parous women aged 18 years or older who were scheduled to undergo a planned induction of labour. Exclusion criteria included the presence of a fetus with a known major structural or genetic anomaly; an abnormal (“Red-Zone”) fetal cardiotocograph as defined by New South Wales (NSW) Health (Appendix 1); fetal breech, brow, or face presentation, clinical suspicion of cephalopelvic disproportion, known or suspected chorioamnionitis, intrapartum haemorrhage exceeding 50 mL, intrapartum pyrexia ≥38 °C, fetal scalp blood sample pH <7.25 or lactate >4 mmol/L, a history of pre-existing maternal diabetes or a history of previous caesarean delivery. These criteria were chosen due to potential safety concerns or perceived high risk of caesarean delivery, a potential primary outcome for a definitive trial.

Transition to active labour was defined as the presence of regular uterine contractions occurring at least every four minutes for a minimum duration of one hour, accompanied by full cervical effacement and cervical dilatation of ≥4 cm, as assessed by the managing midwife or obstetric clinician.

### Data

Baseline characteristics at the time of randomisation were collected from the electronic medical record and included maternal age, parity (number of previous births ≥20 weeks’ gestational age), plurality, self-reported maternal ethnicity, gestational age, gestational diabetes status as per the Australasian Diabetes in Pregnancy Society,^11^ hypertension as defined by the Society of Obstetric Medicine of Australia and New Zealand guidelines (pre-eclampsia, gestational hypertension, essential hypertension, no hypertension),^12^ rationale for induction of labour, need for and method of cervical ripening. Outcomes collected from the electronic medical record included, oxytocin doses, number of episodes of uterine hyperstimulation and tachystole, length of first and second stage of labour, time from randomisation to delivery, mode of delivery, degree of perineal trauma, episiotomy, estimated blood loss, maternal and fetal complications.

Participants were contacted via SMS and telephone at six to eight weeks postpartum and invited to complete an online survey designed to capture participant satisfaction^13^ (also see Trial Outcomes below) and follow-up outcomes. Data were collected electronically and securely stored in a REDCap database.

### Randomisation and Allocation Concealment

Following eligibility confirmation, participants were randomly assigned in a 1:1 ratio to either the intervention or comparator group in random block sizes of four, six or eight, stratified by centre and parity (nulliparous/parous). Randomisation was performed by an on-call study investigator not involved in the participant’s clinical care, using a secure, computer-generated allocation system with concealed assignment to ensure that investigators and clinical staff remained blinded until the appropriate point of clinical management.

### Baseline Induction Procedure

In accordance with standard induction practice across participating sites, all participants commenced labour induction with an infusion of 10 IU of oxytocin in 1 L of normal saline or Hartmann’s solution. This baseline infusion was titrated according to the NSW Health protocol throughout the latent phase of labour.(Appendix 1)

### Intervention

Once active labour and ongoing eligibility were confirmed, the initial oxytocin infusion was discontinued and replaced with the study intervention solution. Participants allocated to the intervention group received 5 IU of oxytocin diluted in 1 L of normal saline or Hartmann’s solution. Infusion rates were titrated in mL/h following the NSW Health oxytocin escalation protocol, ensuring consistent dose adjustments across clinical sites.(Appendix 1)

### Comparator

Participants in the comparator group received the routine oxytocin preparation consisting of 10 IU of oxytocin in 1 L of normal saline or Hartmann’s solution. As with the intervention group, infusion rates were titrated according to the NSW Health protocol.

### Blinding and Unblinding Procedures

Participants and clinicians involved in their care were blinded to group allocation. Oxytocin preparations were identically packaged and labelled with a unique study identification number to maintain allocation concealment. To maintain blinding, the study solution was prepared by two trained members of the research team and/or clinicians not involved with the participant’s direct clinical care. All personnel responsible for preparation were aware of the requirement to maintain blinding.

An investigator or Midwifery Unit Manager (MUM) was available 24 hours per day to facilitate urgent unblinding when clinically indicated.

Unblinding occurred when the following predefined safety or labour-progress criteria were met:

- Participant withdrawal after commencement of the study infusion
- Cervical dilatation <2 cm over a 4-hour period
- Failure to reach full cervical dilatation within 8 hours
- Inadequate uterine contractions during the second stage of labour
- Second stage of labour lasting >3 hours in nulliparous participants or >2 hours in parous participants
- Active pushing lasting >2 hours in nulliparous participants or >1 hour in parous participants
- Clinical concern raised by the treating team regarding maternal or fetal wellbeing
- Completion of two full litres of the blinded oxytocin preparation
- Administration of the blinded preparation at 240 mL/h for >30 minutes without adequate clinical response

If unblinding was required, the responsible investigator or MUM accessed the corresponding study identification number to determine group allocation. The blinded oxytocin infusion was immediately discontinued and replaced with the standard-concentration regimen of 10 IU oxytocin in 1 L of normal saline or Hartmann’s solution. The infusion was initially continued at the same oxytocin delivery rate (IU/min) as the blinded preparation. For participants who had been receiving the lower-dose (5 IU) preparation, the replacement dose was initiated at half the previous infusion rate and escalated thereafter following the NSW Health oxytocin titration protocol.

### Trial outcomes

#### Primary Outcome

The primary outcome of this pilot study was the feasibility of conducting a large multicentre trial. Feasibility was assessed by examining barriers encountered during recruitment, the number of potential participants assessed for eligibility, the proportion consenting to participate, the proportion randomised and the total oxytocin dose and final oxytocin infusion rates to evaluate whether the two treatment allocations resulted in a clinically meaningful difference in oxytocin exposure.

#### Secondary Outcomes

Secondary outcomes were assessed to evaluate maternal and neonatal safety, clinical efficacy, and participant-reported experience. Participant-reported satisfaction with labour and birth experience was measured using a visual analogue scale (VAS) at 1–2 weeks postpartum, with additional questionnaire responses collected at 6–8 weeks postpartum based on the ‘Six simple questions’ questionairre.^13^ (Appendix 2). Clinical outcomes included emergency caesarean delivery, emergency caesarean delivery performed for fetal distress, and emergency operative delivery (forceps or vacuum) for fetal distress. Serious maternal morbidity or mortality, reported as a combined outcome, included postpartum haemorrhage requiring transfusion; third- or fourth-degree perineal trauma; dilatation and curettage for bleeding or retained placental tissue; cervical laceration; vertical uterine incision; vulvar or perineal haematoma; pneumonia; venous thromboembolism requiring anticoagulation; wound infection requiring hospital stay exceeding seven days; readmission for obstetric causes; wound dehiscence; maternal fever ≥38.5°C on two occasions at least 24 hours apart (excluding the first 24 hours); bladder, ureter, or bowel injury requiring repair; genital-tract fistula; bowel obstruction; and intensive care admission. Serious perinatal or neonatal morbidity or mortality within six weeks of birth, also reported as a combined outcome, included shoulder dystocia requiring manoeuvres other than McRoberts or suprapubic pressure or resulting in neonatal injury; 5-minute Apgar scores <4; arterial cord pH <7.0, lactate >10 mmol/L, or base excess <−15; seizures within 24 hours of birth; intubation or ventilation exceeding 24 hours; tube feeding for more than four days; neonatal intensive care unit admission exceeding four days; neonatal jaundice requiring phototherapy; neonatal fracture; intraventricular or intracranial haemorrhage; subgaleal haemorrhage; neonatal blood transfusion; hypoxic-ischaemic encephalopathy (HIE); and neuropraxia.

#### Other Outcomes

Additional outcomes included instrumental birth, fetal distress defined by Red Zone CTG (Appendix 1) or fetal scalp lactate >4.8 mM, fetal acidaemia defined by arterial cord pH <7.1, rates of uterine hyperstimulation, time from randomisation to delivery, duration of the first and second stages of labour, overall duration of labour, estimated blood loss at delivery, second-, third-, or fourth-degree perineal trauma, length of hospital stay, breastfeeding at discharge, and the occurrence of unblinding, both in accordance with the protocol and as protocol deviations. The six to eight week follow-up questions included two questions about participating in a community group to give feedback about the trial and to develop a larger study across hospitals.(Appendix 2)

## Results

Figure 1 shows the participant flowchart. 771 potential participants were assessed for eligibility and 572 were approached Two-hundred and ninety-three (51%) agreed to participate of whom 101 (34%) were randomised. Recruitment ceased between 17^th^ July 2024 and 3^rd^ November 2024 due to an international intravenous fluid shortage.

Of the 199 potential participants deemed ineligible at the initial screening, the most common reasons were non-English speaking, progressed into spontaneous labour, admission under a private obstetrician, and a history of previous caesarean section.

Table 1 shows similar baseline characteristics between the two groups. Table 2 shows primary and secondary outcomes. The median total oxytocin dose per participant was 3.6 IU in the reduced oxytocin group and 7.2 (IU) in the comparison group among participants where blinding was maintained (Wilcoxon rank sum test p <0.0001).(Table 2) The median maximum infusion rates were 10 mIUmin^-1^ in the reduced oxytocin group and 15 mIUmin^-1^ in the comparison group. No differences were observed in any of the clinical outcomes.

**Table 1:** Baseline characteristics among 100 participants.

|  | **Treatment**<br>**(halved oxytocin)**<br>**n=51\*** | **Routine care**<br>**n=50\*** |
| --- | --- | --- |
| Age (years) | 32 (30, 35) | 31 (29, 35) |
| Body mass index (kgm<sup>-2</sup>) | 23.7 (21.6, 26.2) | 25.2 (21.3, 27.9) |
| Gestational age (weeks+days) | 39<sup>+4</sup> (39<sup>+1</sup>, 40<sup>+2</sup>) | 39<sup>+5</sup> (39<sup>+1</sup>, 40<sup>+2</sup>) |
| Nulliparous | 41 (80) | 41 (82) |
| Plurality |  |  |
| Singleton | 50 (98) | 50 (100) |
| Twins | 1 (2) | 0 (0) |
| Place of origin |  |  |
| Australia/Oceania | 22 (43) | 18 (36) |
| East/central Asia | 3 (6) | 5 (10) |
| South/South East Asia | 13 (25) | 17 (34) |
| Europe | 6 (12) | 6 (12) |
| Middle East | 3 (6) | 0 (0) |
| Other | 4 (8) | 4 (8) |
| Royal Prince Alfred | 46 (90) | 46 (92) |
| Canterbury | 5 (10) | 4 (8) |
| Indication for induction of labour |  |  |
| Decreased fetal movements | 12 (24) | 7 (14) |
| Gestational diabetes | 9 (18) | 10 (20) |
| Post-term | 8 (16) | 7 (14) |
| Suspected fetal compromise | 7 (14) | 3 (6) |
| Suspected large fetus | 7 (14) | 2 (4) |
| Prelabour ruptured membranes | 0 (0) | 5 (10) |
| Hypertension in pregnancy | 0 (0) | 4 (8) |
| Cholestasis of pregnancy | 0 (0) | 2 (4) |
| Other** | 8 (16) | 10 (20) |
| Diabetes in pregnancy |  |  |
| None | 42 (82) | 38 (76) |
| Treated with diet | 3 (6) | 8 (16) |
| Treated with diet and insulin | 6 (12) | 4 (8) |
| Hypertension in pregnancy |  |  |
| None | 51 (100) | 46 (92) |
| Essential hypertension | 0 (0) | 2 (4) |
| Gestational hypertension | 0 (0) | 1 (2) |
| Pre-eclampsia | 0 (0) | 1 (2) |
| Cervical ripening | 46 (90) | 40 (80) |
| Cervical ripening method |  |  |
| None | 5 (10) | 10 (20) |
| Balloon | 3 (6) | 2 (4) |
| Prostglandins | 24 (47) | 21 (42) |
| Both | 19 (37) | 17 (34) |
| Pre-labour artificial rupture of membranes | 39 (76) | 37 (74) |
| Epidural analgesia before randomisation | 25 (49) | 28 (56) |

\* n (%) or median (interquartile range)
\*\*Other indications for induction of labour included advanced maternal age, dichorionic twins, in-vitro fertilisation, fetal anomaly, prolonged latent phase of the first stage of labour, ethnicity, thrombocytopaenia, and combinations of risk factors.

**Table 2:** Primary and secondary outcomes.

|  | **Treatment**<br>**(halved**<br>**oxytocin)**<br>**n=51\*** | **Routine care**<br>**n=50\*** | **p\*** |
| --- | --- | --- | --- |
| Total oxytocin dose (IU)** | 3.6 (2.0, 4.9) | 7.2 (3.0, 11.7) | <0.0001 |
| Maximum oxytocin infusion rate<br>(mU/min)** | 10 (7.5, 12.5) | 15 (10, 25) | <0.0001 |
| Caesarean delivery | 10 (20) | 13 (26) | 0.44 |
| Caesarean delivery for fetal concerns | 4 (8) | 2 (4) | 0.68 |
| Operative delivery for fetal concerns | 20 (39) | 19 (38) | 0.90 |
| Serious maternal morbidity or mortality | 2 (4) | 3 (6) | 1.0 |
| Serious perinatal morbidity or mortality | 10 (20) | 6 (12) | 0.28 |

\* n (%) or median (interquartile range); Wilcoxon rank sum test, X<sup>2</sup> test, or Fisher's exact test
\*\* among 45 (treatment group) and 40 (routine care group) where allocation remained unblinded

Table 3 shows results for the other outcomes. Clinical and satisfaction outcomes were similar between the two study groups. Satisfaction outcomes were collected for 69% (70/101) participants. There was 100% ascertainment for short-term clinical outcomes and for readmissions to the maternity hospital of delivery. Six-to-eight-week follow-up was obtained for 65 participants.

**Table 3:** Other Outcomes.

|  | **Treatment**<br>**(halved oxytocin)**<br>**n=51\*** | **Routine care**<br>**n=50\*** | **p\*** |
| --- | --- | --- | --- |
| Satisfaction with labour and birth experience (VAS) | 9 (8, 10)<br>n=36 | 8 (7, 9.75)<br>n=34 | 0.22 |
| Would choose again<br>(1=strongly disagree; 7=strongly agree) | 7 (5.75, 7)<br>n=36 | 7 (6, 7)<br>n=34 | 0.62 |
| Instrumental birth | 22/51 (43) | 20/50 (40) | 0.75 |
| 'Fetal distress' | 30/50 (60) | 26/50 (52) | 0.42 |
| Fetal acidaemia | 15/50 (30) | 12/50 (24) | 0.50 |
| Uterine tachysystole |  |  |  |
| No episodes | 45 (90) | 47 (94) | 0.31 |
| 1 episode | 5 (10) | 2 (4) |  |
| 2 episodes | 0 (0) | 1 (2) |  |
| Uterine hypertonus |  |  |  |
| No episodes | 43 (86) | 41 (82) | 0.57 |
| 1 episode | 7 (14) | 8 (16) |  |
| 2 episodes | 0 (0) | 1 (2) |  |
| Time from randomisation to delivery (h) | 6.1 (4.1, 8.8) | 6.0 (4.4, 8.1) | 0.98 |
| Length of first stage of labour (h) | 5.0 (4.1, 8.2) | 5.1 (3.8, 7.9) | 0.83 |
| Length of second stage of labour (h) | 2.0 (0.8, 3.3) | 2.1 (1.2, 2.9) | 0.90 |
| Overall duration of labour (h) | 7.2 (4.4, 10.3) | 6.6 (5.2, 8.3) | 0.76 |
| Estimated blood loss at delivery (mL) | 450 (300, 800) | 400 (300, 650) | 0.61 |
| Perineal trauma |  |  |  |
| None or 1<sup>st</sup> degree | 26 (51) | 22 (44) | 1.0** |
| 2<sup>nd</sup> degree | 25 (49) | 27 (54) |  |
| 3<sup>rd</sup> or 4<sup>th</sup> degree | 0 (0) | 1 (2) |  |
| Length of hospital stay (days) | 4.10 (3.22, 5.28) | 4.58 (3.29, 5.47) | 0.43 |
| Exclusive breast feeding at discharge | 26/50 (52) | 27/50 (54) | 0.84 |
| Unblinding | 6/50 (12) | 9/51 (18) | 0.42 |
| In accordance with protocol | 3 | 9 |  |
| Not in accordance with protocol | 3 | 1 |  |

\* n (%) or median (interquartile range); Wilcoxon rank sum test, X<sup>2</sup> test, or Fisher's exact test
\*\* For 3<sup>rd</sup>/4<sup>th</sup> degree versus others
VAS = visual analogue scale

Table 4 shows rates of recruitment. Recruitment occurred at a rate of 9.2% (62/1205) of all births when a research midwife was employed.

**Table 4:** Proportion of all births that were recruited to the EASE-OUT Feasibility Trial.

| **Research Midwife** | **Hospital** | **Dates** | **Days/ week** | **Total days** | **Births in the period** | **n\*** | **Randomisations per birth** |
| --- | --- | --- | --- | --- | --- | --- | --- |
| 1 | RPA | Sept – Dec 2023 | 2 | 20 | 230 | 24 | 10.4% |
| None^ | RPA | Dec 2023 - Mar 2024 | 7 | 78 | 897 | 18 | 2.0% |
| 2 | RPA | Mar – July 2024** | 2 | 32 | 368 | 29 | 7.9% |
| IV fluid shortage: recruitment ceased 17<sup>th</sup> July– 3<sup>rd</sup> November 2024 |  |  |  |  |  |  |  |
| 3 | Canterbury Hospital | Jan – Jun 2025& | 1 | 19 | 78 | 9 | 11.5% |

\*n = number of randomisations in the time-period
\*\*2 weeks leave excluded
^A research midwife was not employed to recruit over this period
&5 weeks leave excluded
RPA = Royal Prince Alfred Hospital

At the six to eight week follow-up, 29 participants indicated they were interested in being part of a community group to provide feedback about the study and to be involved in developing a larger study across multiple hospitals, and 28 participants agreed to receive more information about this.

## Discussion

Our main finding was that large blinded randomised controlled trials of oxytocin management in labour are feasible. We were able to approach 572 potential participants in two centres for a consent rate of 51%, with 9.2% of all births in the participating hospitals being randomised to the EASE-OUT Feasibility Trial during periods of active recruitment. We also found that a lower median total dose of oxytocin was required (7.2 IU v 3.6 IU; p <0.0001) when the rate of infusion was halved compared with usual care and that the median maximum infusion rate was lower by 5 mIUmin^-1^ (10 v 15 mIUmin^-1^; p <0.0001). These differences support the idea that halving the rate of oxytocin could be tested in a randomised controlled trial, potentially overcoming some of the barriers to implementing complete cessation of the medication,^9,14^ and that there is sufficient treatment differentiation between the two treatments to impact on outcomes. Further, our follow-up finding that 29 participants were interested in providing feedback about the trial and being involved in developing a larger study across multiple hospitals indicates it would be feasible to find community members to advise and collaborate on a multicentre trial.

These results are important for assessing feasibility. We found that eligible participants were receptive to participation in an intrapartum trial with more than 50% of those approached agreeing to participate, and that a larger trial is feasible. For a theoretical sample size of 1695 for the outcome of caesarean delivery (α 0.05, β 0.10, baseline, rate 25%, clinically important reduction to 20%), such a trial would require ten participating hospitals with 3300 annual births each and a recruitment phase of one year. This estimation conservatively assumes recruiting 5% of all births (compared with 9.2% observed in this feasibility trial) and accounts for observed rates of not approaching eligible participants, rates of declining to participate, and missing randomisation for some consented participants.

There is strong evidence that lowering oxytocin doses can impact on uterine hyperstimulation without increasing the rate of caesarean section. Ceasing oxytocin infusions in the active phase of induced labours reduces uterine tachsystole and abnormal fetal heart rate traces in labour, and caesarean deliveries may also be reduced.^8^ However, there are significant barriers to compliance including impatience with slower than average rates of cervical dilatation, pressure from colleagues to restart the oxytocin, and preconceived ideas that ceasing oxytocin leads to higher rates of caesarean delivery.^9,14^ Thus, it is important to assess alternative, more acceptable methods of reducing oxytocin, with appropriate implementation strategies.

While induction of labour at term is associated with generally improved outcomes for mothers and babies,^15-17^ oxytocin is associated with fetal distress due to umbilical cord compression, and abnormal fetal heart rate traces are in turn associated with HIE and long-term neurological dysfunction.^18-20^ Reducing uterine over-stimulation through alternative regimens of oxytocin administration could further increase the clinical benefits of term induction of labour. Other options include limiting the frequency with which oxytocin infusions can be increased (e.g., hourly or 2-hourly), targeting different frequencies of contractions (e.g., three per 10 minutes versus 4 per ten minutes), or halving the dose of oxytocin in active labour and maintaining that rate for two hours or more before titrating upwards. These options, and others, are particularly suited to an adaptive platform trial where multiple interventions can be assessed more efficiently than in a traditional randomised controlled trial.^21^ Any proposed trial needs to consider the satisfaction with care and experiences of labour and childbirth, both in the planning and as important outcomes.

Strengths of the EASE-OUT Feasibility Trial include its randomised design and feasibility outcomes. Blinding of the oxytocin preparations is expected to reduce bias. The focus on feasibility outcomes allowed identification of causes for protocol deviations and unblinding, which will be invaluable for a larger study as it will help refine the eligibility criteria, escalation pathways and identify key areas for staff training.

The main limitations were the loss to follow up after six weeks, which can lead to selection bias for the longer-term outcomes. There is also a possibility that blinding may be compromised due to observations around the frequency and strength of contractions and we were unable to assess the extent to which this may have occurred. Aas a feasibility trial, this study had insufficient sample size to assess clinical outcomes.

Further research should focus on identifying safer, effective methods of using oxytocin to induce labour, aiming to strike the right balance between adequate progress in labour and the complications of over-stimulating the uterus which include fetal distress, intrapartum stillbirth and HIE. A future definitive trial remains justified. Future studies should focus on identifying an oxytocin reduction strategy that is acceptable to clinicians, practical within labour ward workflows, and capable of reducing uterine overstimulation without compromising labour progress or increasing operative birth. Most importantly, any future trials need to account for and measure birth experiences and have substantial community involvement.

## CONCLUSION

This multicentre, double-blinded feasibility trial demonstrated the feasibility of a randomised trial evaluating oxytocin dose reduction during active labour, with successful recruitment, randomisation, intervention implementation and participant follow-up.

## Data Availability

All data produced in the present study are available upon reasonable request to the authors and approval by Sydney Local Health Ethics Committee.

## Acknowledgements

We would like to thank the Sydney Institute for Women, Children and their Families (SIWCF) and the NHMRC Clinical Trials Centre for providing seed funding; Pejman Adily, database manager for the SIWCF, for creating and uploading the randomisation schedule; the Midwifery Unit Managers, midwives and doctors for their time spent supporting the trial, and the members of the data safety monitoring board for providing clinical and safety oversight.

## Appendix 1

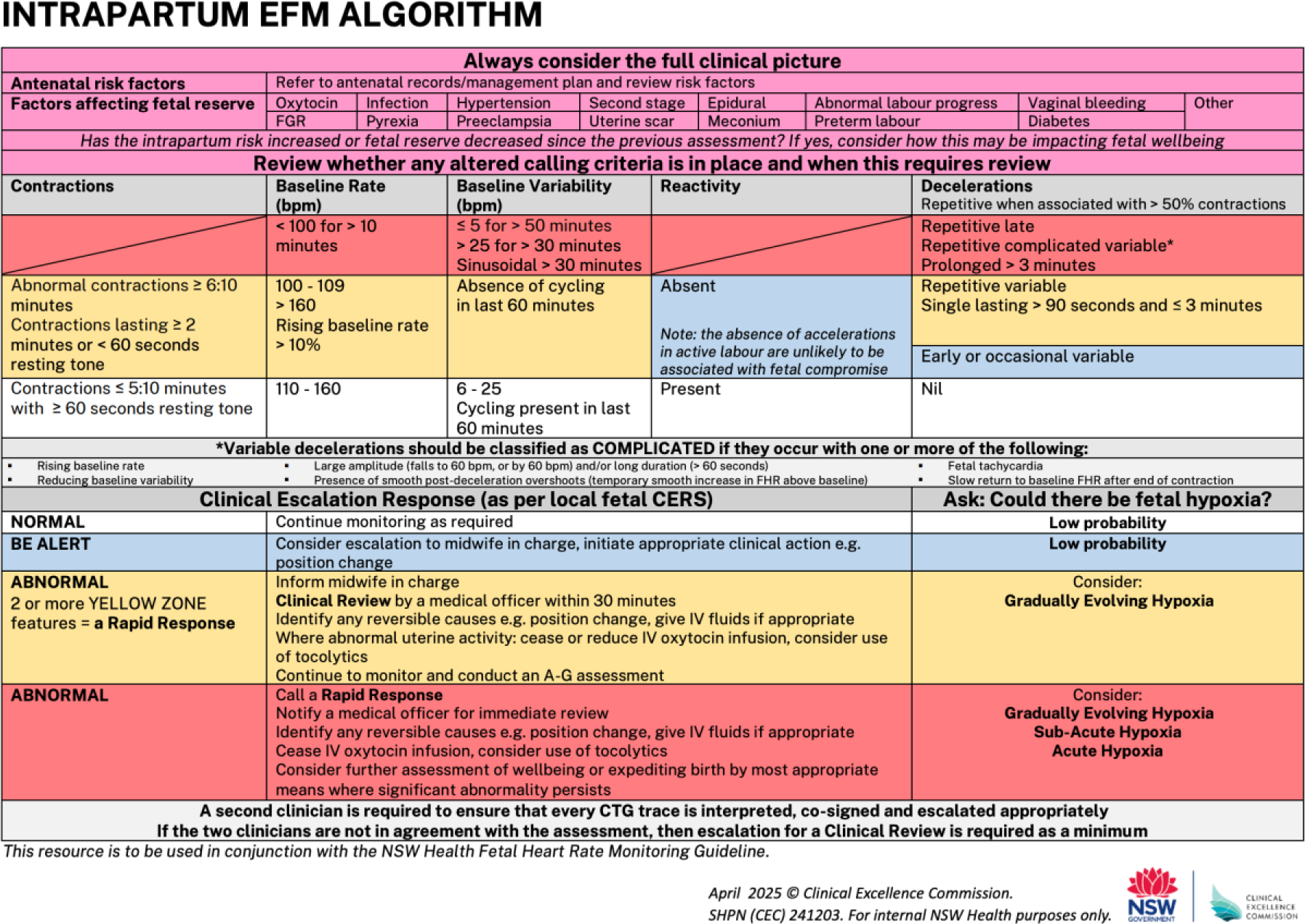

### NSW Health Oxytocin protocol, current during the EASE-OUT Feasibility Trial

| Time | Milliunits per minute | MLs per hour |
| --- | --- | --- |
| Start | 2.5 | 15 |
| Min 30 mins | 5 | 30 |
| ½ hourly | 10 | 60 |
|  | 15 | 90 |
|  | 20 | 120 |
|  | 25 | 150 |
|  | 30 | 180 |
| Medical Review |  |  |
| ½ hourly | 35 | 210 |
|  | 40 | 240 |

## Appendix 2

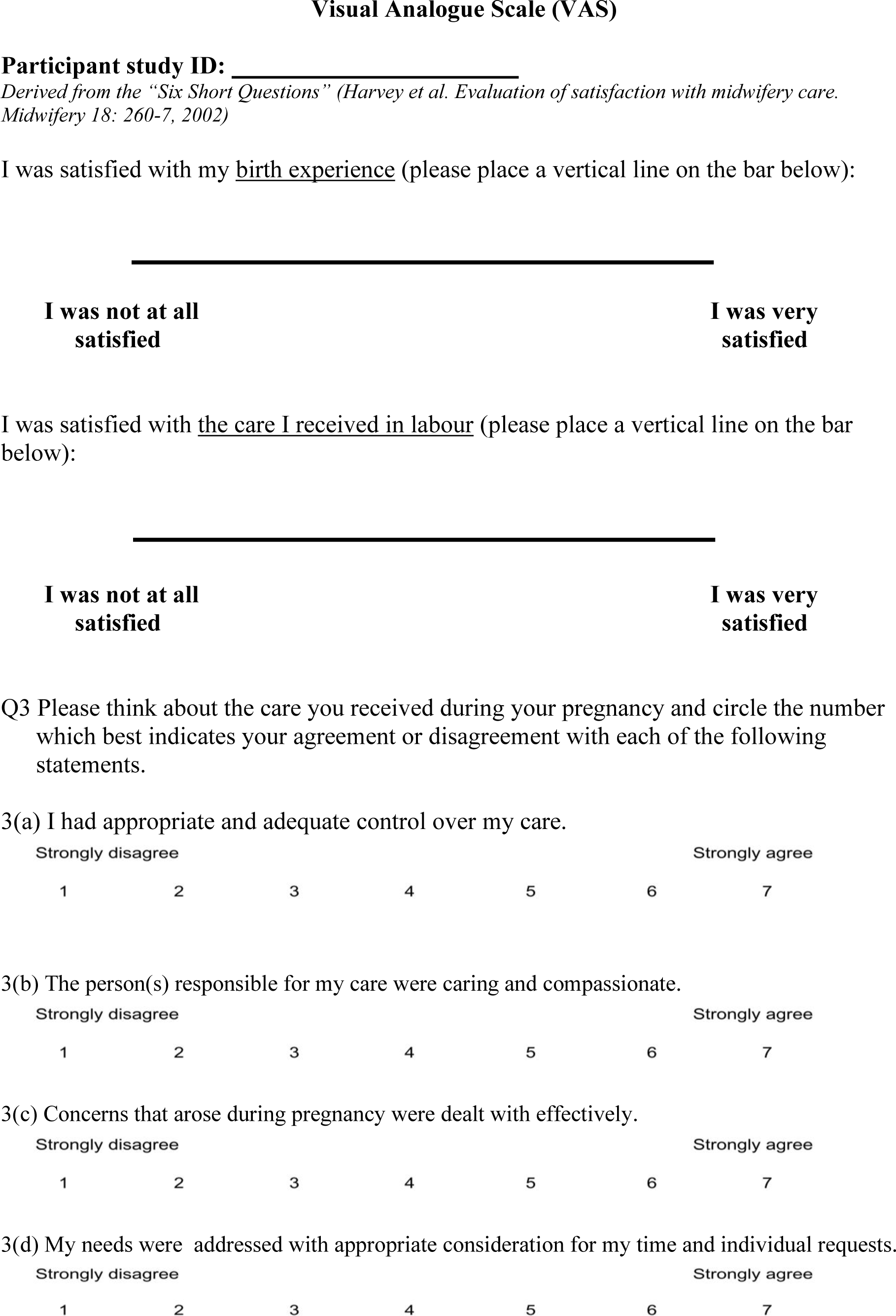
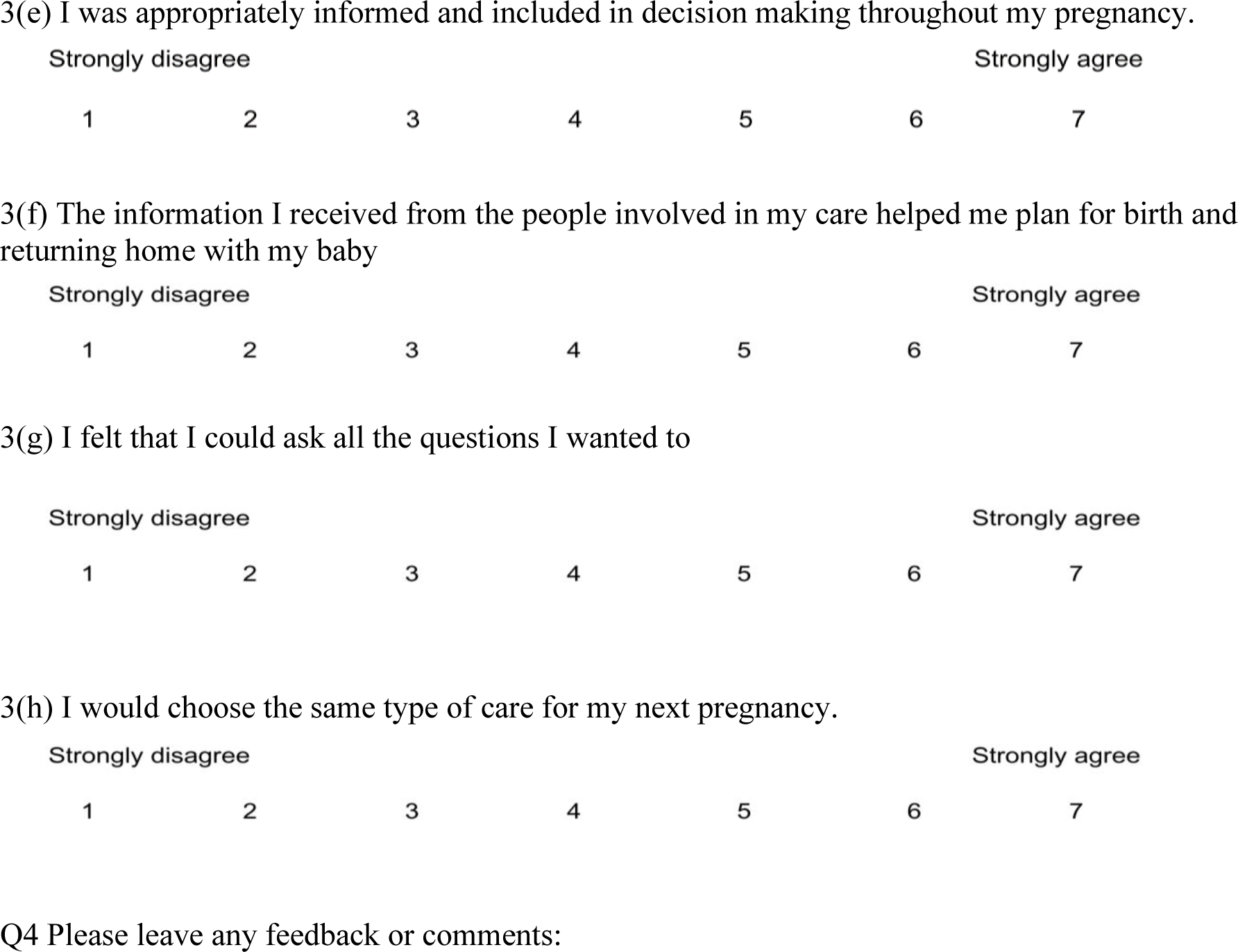

### Follow up questions at 6-8 weeks after birth

- Have you been well since your birth?
- If not, in what way have you been unwell?
- Have you had twins?
- Has your baby been well since it was born?
- Has twin 1 been well since it was born?
- If no, in what way has your baby been unwell?
- Has twin 2 been well since it was born?
- If no, in what way has your baby been unwell?
- Have you been back to hospital or seen a doctor for any reason since your baby was born?
- If yes, for what reason?
- Has your baby been back to hospital or seen a doctor for any reason since they were born?
- If yes, for what reason?
- What method of feeding your baby are you engaging in?
- What other feeding?

### Two community participation questions

- Would you be interested in being a part of community group to provide feedback about this study and to be involved in developing a larger study across multiple hospitals?
- Would it be okay if we sent you some more information about this and how to contact us if you are still interested?

## Notes

### Competing Interest Statement

The authors have declared no competing interest.

### Clinical Trial

ACTRN12622001342707

### Author Declarations

The study protocol underwent review and received formal approval from the Ethics Review Committee of the Sydney Local Health District prior to commencement of participant recruitment (Approval No. X22-0294).

## References

1. Australian Institute of Health and Well-being. Australia’s mothers and babies. Web report | Last updated: 30 Jun 2026 (accessed at https://www.aihw.gov.au/reports/mothers-babies/australias-mothers-babies/contents/labour-and-birth/onset-of-labour on 16th July 2026).

2. Nicholson SM, Hatt S, Kent EM, et al. The rising tide: Trends in induction of labor at term over a 5-year period at a single centre. Int J Gynaecol Obstet 2025; 169(1): 383–90.

3. Martin JA, Osterman MJK. Induction of Labor Increases in the United States: 2016 to 2024. NCHS Data Brief 2026; (554).

4. Swift EM, Gunnarsdottir J, Zoega H, Bjarnadottir RI, Steingrimsdottir T, Einarsdottir K. Trends in labor induction indications: A 20-year population-based study. Acta Obstet Gynecol Scand 2022; 101(12): 1422–30.

5. Brahmawar Mohan S, Sommerfelt H, Froen JF, et al. Antenatal Uterotonics as a Risk Factor for Intrapartum Stillbirth and First-day Death in Haryana, India: A Nested Case-control Study. Epidemiology 2020; 31(5): 668-76.

6. Burgod C, Pant S, Morales MM, et al. Effect of intra-partum Oxytocin on neonatal encephalopathy: a systematic review and meta-analysis. BMC Pregnancy Childbirth 2021; 21(1): 736.

7. Simpson KR, James DC. Effects of oxytocin-induced uterine hyperstimulation during labor on fetal oxygen status and fetal heart rate patterns. Am J Obstet Gynecol 2008; 199(1): 34 e1-5.

8. Whitley J, Burd J, Doering M, Kelly J, Frolova A, Raghuraman N. Reduced risk of cesarean delivery with oxytocin discontinuation in active labor: a systematic review and meta-analysis. Am J Obstet Gynecol 2025; 233(1): 25–39 e11.

9. O’Dea S, Davis G, Balendran J, Phipps H, O’Brien K, de Vries B. Ceasing oxytocin in the active phase of the first stage of induced labours: A prospective audit at a tertiary hospital. medRxiv (preprint): DOI pending 2026.

10. Budden A, Chen LJ, Henry A. High-dose versus low-dose oxytocin infusion regimens for induction of labour at term. Cochrane Database Syst Rev 2014; (10): CD009701.

11. Nankervis A, McIntyre HD, Moses R, et al. ADIPS Consensus Guidelines for the Testing and Diagnosis of Gestational Diabetes Mellitus in Australia. Accessible at: http://adips.org/downloads/ADIPSConsensusGuidelinesGDM-03.05.13VersionACCEPTEDFINAL.pdf. 2013.

12. Lowe SA, Bowyer L, Lust K, et al. The SOMANZ Guidelines for the Management of Hypertensive Disorders of Pregnancy 2014. Aust N Z J Obstet Gynaecol 2015; 55(1): 11–6.

13. Harvey S, Rach D, Stainton MC, Jarrell J, Brant R. Evaluation of satisfaction with midwifery care. Midwifery 2002; 18(4): 260–7.

14. Boie S, Glavind J, Uldbjerg N, Steer PJ, Bor P, group Ct. Continued versus discontinued oxytocin stimulation in the active phase of labour (CONDISOX): double blind randomised controlled trial. BMJ 2021; 373: n716.

15. de Vries BS, Gordon AG. Induction of labour at 39 weeks should be routinely offered to low-risk women. ANZJOG Accepted for publication 2019.

16. Middleton P, Shepherd E, Morris J, Crowther CA, Gomersall JC. Induction of labour at or beyond 37 weeks’ gestation. Cochrane Database Syst Rev 2020; 7: CD004945.

17. Sotiriadis A, Petousis S, Thilaganathan B, et al. Maternal and perinatal outcomes after elective induction of labor at 39 weeks in uncomplicated singleton pregnancy: a meta-analysis. Ultrasound Obstet Gynecol 2019; 53(1): 26–35.

18. Turner J, Kumar S. Neurodevelopmental outcomes in infants following intrapartum maternal oral sildenafil citrate treatment. Am J Obstet Gynecol 2021; 224(3): 316–7.

19. Chakkarapani E, de Vries LS, Ferriero DM, Gunn AJ. Neonatal encephalopathy and hypoxic-ischemic encephalopathy: the state of the art. Pediatr Res 2025; 98(7): 2444–58.

20. Eenkhoorn C, van den Wildenberg S, Goos TG, Dankelman J, Franx A, Eggink AJ. A systematic catalog of studies on fetal heart rate pattern and neonatal outcome variables. J Perinat Med 2025; 53(1): 94–109.

21. Berry SM, Connor JT, Lewis RJ. The platform trial: an efficient strategy for evaluating multiple treatments. JAMA 2015; 313(16): 1619–20.

